# SARS-CoV-2-specific peripheral T follicular helper cells correlate with neutralizing antibodies and increase during convalescence

**DOI:** 10.1101/2020.10.07.20208488

**Authors:** Sushma Boppana, Kai Qin, Jacob K Files, Ronnie M. Russell, Regina Stoltz, Frederic Bibollet-Ruche, Anju Bansal, Nathan Erdmann, Beatrice H. Hahn, Paul Goepfert

**Author notes:** Corresponding author: Paul Goepfert, Bevill Biomedical Research Building Rm 563, 845 19^th^ Street S, Birmingham, AL 35294, USA. These authors contributed equally (Sushma Boppana, Kai Qin).

## Abstract

T-cell immunity is likely to play a role in protection against SARS-CoV-2 by helping generate neutralizing antibodies. We longitudinally studied CD4 T-cell responses to the M, N, and S structural proteins of SARS-CoV-2 in 21 convalescent individuals. Within the first two months following symptom onset, a majority of individuals (81%) mount at least one CD4 T-cell response, and 48% of individuals mount detectable SARS-CoV-2-specific peripheral T follicular helper cells (pTfh, defined as CXCR5^+^PD1^+^ CD4 T cells). SARS-CoV-2-specific pTfh responses across all three protein specificities correlate with antibody neutralization with the strongest correlation observed for S protein-specific responses. When examined over time, pTfh responses increase in frequency and magnitude in convalescence, and robust responses with magnitudes greater than 5% were detected only at the second convalescent visit, an average of 38 days post-symptom onset. These data deepen our understanding of antigen-specific pTfh responses in SARS-CoV-2 infection, suggesting that M and N protein-specific pTfh may also assist in the development of neutralizing antibodies and that pTfh response formation may be delayed in SARS-CoV-2 infection.

**Author Summary:** Since December 2019, the Coronavirus Disease 2019 (COVID-19) pandemic has caused significant morbidity and mortality worldwide. Most currently licensed vaccines are understood to protect against infection by inducing neutralizing antibodies. As such, ongoing COVID-19 vaccine trials have focused on antibody neutralization as a primary immunologic endpoint. It is well established that T follicular helper cells are essential to the development of neutralizing antibodies and that a subset of these cells, peripheral T follicular helper cells (pTfh), can be studied in the blood. However, little is known about Tfh responses mounted in SARS-CoV-2 infection. Here, we studied pTfh to three major structural proteins in individuals recovered from COVID-19. We find that SARS-CoV-2-specific pTfh frequencies correlate with neutralizing antibody responses, especially those directed against the spike protein. We also find that pTfh responses to SARS-CoV-2 increase over time. Our findings suggest that pTfh responses against proteins other than the spike protein may contribute to the development of neutralizing antibodies and suggests that formation of pTfh responses in SARS-CoV-2 infection may be delayed.

## Introduction

Cases of COVID-19, caused by the novel severe acute respiratory syndrome coronavirus 2 (SARS-CoV-2), were first reported in Wuhan, China at the end of 2019 (1). Since then, the COVID-19 pandemic has caused significant morbidity, mortality, and economic disruption worldwide (2). In SARS-CoV-2 infection, initial studies reported significant lymphopenia in hospitalized patients (3). An elevation of both activation and exhaustion markers on T cells in both severe and mild disease has also been described (4-6). More recently, data on antigen-specific T-cell responses in individuals recovered from SARS-CoV-2 infection has emerged. These studies have reported CD4 T-cell responses to SARS-CoV-2 in 80-100% of convalescent individuals, with most publications focusing on the Spike (S) protein (7-10).

Several SARS-CoV-2 vaccine efficacy trials are in progress, and recent Phase I/II trial data have highlighted the presence of neutralizing antibodies as evidence of plausible vaccine efficacy (11-13). Although the key components of a protective immune response against SARS-CoV-2 remain unclear, studies in non-human primates have found that neutralizing antibodies (nAb) are a correlate of protection in infection and vaccination (14, 15). With this in mind, a better understanding of how T-cell responses contribute to the formation of nAb is critical to optimizing future vaccine design.

Because direct study of lymphoid tissues in humans is difficult, peripheral T follicular cells (pTfh), or T follicular helper cells (Tfh) circulating in the blood, serve as an important surrogate for understanding Tfh responses within germinal centers. While there is some controversy regarding how to best identify these cells, there is general consensus that these cells express CXCR5, a lymph node homing receptor, and many groups use PD1 expression in conjunction with CXCR5 to define pTfh (16-18). While frequencies of circulating CXCR5^+^PD1^+^ CD4 T cells are typically low, these cells are closely linked to Tfh in lymphoid tissue (19) and have been shown to support humoral responses (20, 21). Antigen-specific pTfh have been shown to correlate with neutralizing antibodies in the context of infection and vaccination of several pathogens (17, 22-26). Although pTfh responses have not been described in the context of SARS-CoV or MERS-CoV infection, CD4 T-cell responses have been shown to be important in controlling SARS-CoV in mouse models (27), and a recent study of a MERS-CoV vaccine in mice found that Tfh frequencies in draining lymph nodes correlated with neutralizing antibodies (28).

Data on SARS-CoV-2-specific T follicular helper cell responses are also limited. Thevarajan et al. was the first to report on pTfh frequencies in SARS-CoV-2, and found that frequencies of total pTfh increased during acute infection (29). Since then, a few studies have drawn a correlation between total CD4 T cell or total Tfh-like cell frequencies and antibody levels (30, 31). Another study found increased expression of CXCR5 and ICOS, two Tfh markers, on SARS-CoV-2-specific CD4 T-cells but did not examine pTfh responses directly (32). In deceased donors with COVID-19, Kaneko et al. recently reported that BCL6-expression in germinal center Tfh was lost within thoracic lymph nodes. This study suggests that Tfh response formation may be impaired in severe SARS-CoV-2 infection (33), but how this affects the formation of antigen specific Tfh responses is unclear.

The most direct examination of pTfh to date was conducted by Juno et al, where circulating Tfh in the blood were defined as CD45RA^-^CXCR5^+^ CD4 T cells. They demonstrated a correlation between S protein-specific pTfh and nAb, suggesting that Tfh responses are formed in mild SARS-CoV-2 infection (34). However, these data leave several questions unanswered, including at what point in convalescence these responses evolve and whether Tfh specific for other SARS-CoV-2 proteins contribute to the formation of neutralizing antibodies. While this study was a useful first glimpse at antigen-specific Tfh responses, it did not examine PD1 expression, a canonical Tfh marker, and used the activation markers, Ox40 and CD25, to identify antigen-specific responses, which have previously been shown include a high percentage of T regulatory cells (35). It is also important to note that pTfh specificity does not necessarily correspond with neutralizing antibody specificity. For example, in HIV infection and vaccination, intrastructural help occurs, where CD4 T-cell responses to internal, structural proteins correlated with neutralizing antibodies against the exterior, envelope protein (36, 37). These studies underscore the importance of examining pTfh responses across the SARS-CoV-2 proteome.

Here, we report on SARS-CoV-2-specific CD4 T-cell responses to the membrane (M), nucleocapsid (N), and spike (S) proteins studied longitudinally in 21 convalescent individuals. We directly examined antigen-specific pTfh (CXCR5^+^PD1^+^ CD4 T cells) and observed correlations between antigen-specific pTfh responses across all protein specificities and antibody neutralization, with the strongest correlation observed for S protein-specific pTfh frequencies. High magnitude SARS-CoV-2-specific pTfh responses (>5% activation of total pTfh population) were only detected at the second convalescent visit, more than 30 days following symptom onset. These data are the first to examine the kinetics of pTfh responses that arise after SARS-CoV-2 infection as well as the relationship between neutralizing antibodies and pTfh responses to the SARS-CoV-2 M and N proteins. These results also suggest that pTfh formation may be delayed in SARS-CoV-2 infection.

## Results

### SARS-CoV-2-specific CD4 T cells target the M, N, and S proteins in individuals recovered from COVID-19 at their first convalescent visit

In 21 individuals recovered from COVID-19, we assessed the presence of T-cell responses to the membrane (M), nucleocapsid (N), and spike (S) proteins of SARS-CoV-2 using overlapping 20mer peptide pools spanning each protein. All but two of these individuals were confirmed to have SARS-CoV-2 infection by PCR, and the two who were not PCR tested had a known COVID-19 contact and detectable SARS-CoV-2-specific T-cell responses. While none of these individuals required hospitalization, all experienced COVID-19 related symptoms, and a majority (71%) reported a moderate severity of symptoms. T-cell responses were measured at the first convalescent visit for each individual, which occurred an average of 22 days post-symptom onset while the second visit was an average of 39 days post-symptom onset (**Table 1**). We utilized two flow cytometry-based strategies: 1) upregulation of activation-induced markers (AIM), and 2) production of effector molecules by intracellular cytokine staining (ICS). Gating strategies for AIM and ICS in an unstimulated, negative control are shown in **S1 Fig**.

**Table 1:**
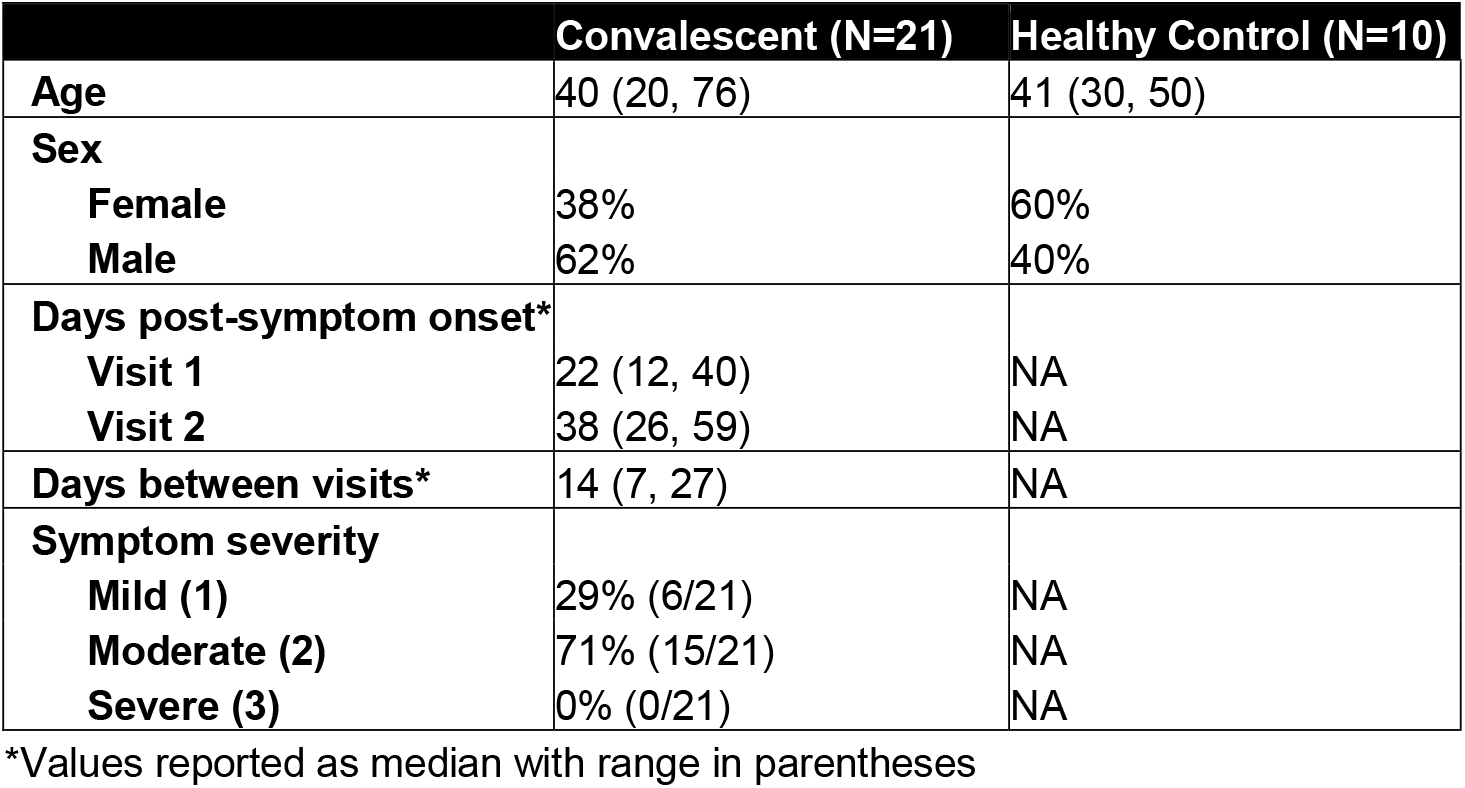
Patient demographics.

|  | Convalescent (N=21) | Healthy Control (N=10) |
| --- | --- | --- |
| **Age** | 40 (20, 76) | 41 (30, 50) |
| **Sex** |  |  |
| **Female** | 38% | 60% |
| **Male** | 62% | 40% |
| **Days post-symptom onset\*** |  |  |
| **Visit 1** | 22 (12, 40) | NA |
| **Visit 2** | 38 (26, 59) | NA |
| **Days between visits\*** | 14 (7, 27) | NA |
| **Symptom severity** |  |  |
| **Mild (1)** | 29% (6/21) | NA |
| **Moderate (2)** | 71% (15/21) | NA |
| **Severe (3)** | 0% (0/21) | NA |

\*Values reported as median with range in parentheses

Representative positive CD4 T-cell responses measured by each staining strategy are shown in **Fig 1A** for AIM and in **Fig 1B** for ICS in one individual, CR8, who mounted CD4 T-cell responses against all three SARS-CoV-2 proteins. At the first convalescent visit, we found that 57% (12/21) of individuals mounted a SARS-CoV-2-specific CD4 T-cell response by AIM and that these CD4 responses targeted all three tested proteins with similar frequencies (**Fig 1C**). Meanwhile, by ICS, 47% (10/21) of individuals had at least one SARS-CoV-2-specific CD4 response at this visit, with a similar distribution across the tested proteins (**Fig 1D**). As a control, we also measured T cell responses to SARS-CoV-2 peptide pools in COVID-19 negative individuals by assaying samples collected from healthy individuals before the COVID-19 pandemic. In the healthy controls tested, we detected three low magnitude (≤0.17%), presumably cross-reactive memory CD4 T-cell responses in two of the ten tested individuals (20%) in line with previously published reports (8). Representative staining of an AIM-detected and an ICS-detected response in healthy controls is shown in **S2A-B Fig**, with overall responder frequencies presented in **S2C-D Fig**. Overall, our data show that nearly half of convalescent individuals mounted a SARS-CoV-2 specific CD4 T-cell response as detected by both activation marker expression and cytokine production.

**Figure 1:**
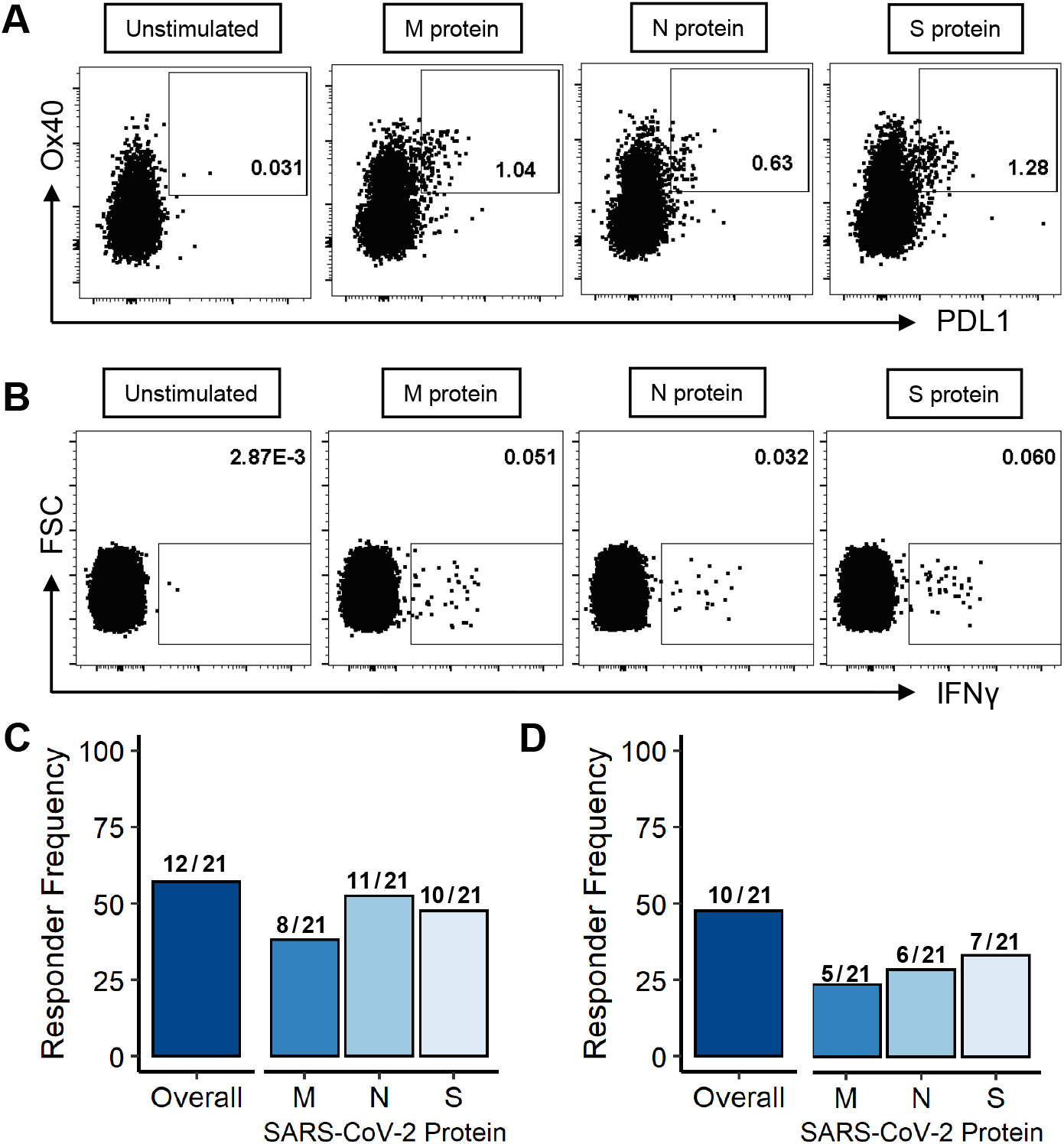
SARS-CoV-2-specific CD4 T cells target the M, N, and S proteins in individuals recovered from COVID-19 at their first convalescent visit. Representative examples of CD4 responses in CR8 to the M, N, and S protein peptide pools as detected by upregulation of activation-induced markers, Ox40 and PDL1 **(A)** and by IFNγ in intracellular cytokine staining **(B)**. Responder frequency of CD4 responses to any SARS-CoV-2 protein and to the M, N, and S proteins individually by AIM **(C)** and ICS **(D)**.

While there was a weak correlation between the response magnitude for AIM and ICS for each condition (**S3A Fig**), more responses were identified by upregulation of activation-induced marker expression than by intracellular cytokine staining. There were 12 responses detected by only AIM that were not positive on ICS, but only one response was detected by ICS only (**S3B Fig**). These data show that a significant portion of CD4 responses detected in early convalescence were not detected by cytokine (IFNγ, TNFα, or CD154) staining and highlight the increased sensitivity of AIM for detecting total CD4 T-cell responses.

### SARS-CoV-2-specific peripheral T follicular helper cells are detected in convalescent individuals

We directly measured antigen-specific pTfh responses by the upregulation of Ox40 and PDL1 on CXCR5^+^PD1^+^ CD4 T cells (gating strategy shown in **S1 Fig**). Representative examples of SARS-CoV-2-specific pTfh responses to the M, N, and S proteins are shown in **Fig 2A**. At the first convalescent visit, occurring an average of 23 days post-symptom onset, we detected 6 total pTfh responses in 4 of the 21 individuals tested (19%) and equally spread across each of the three proteins (**Fig 2B**). These data indicate that only a minority of individuals have mounted detectable SARS-CoV-2-specific pTfh responses early in convalescence. However, previous studies on pTfh responses have rarely calculated responder rates, and, therefore, it is difficult to conclude whether this responder frequency is atypical.

**Figure 2:**
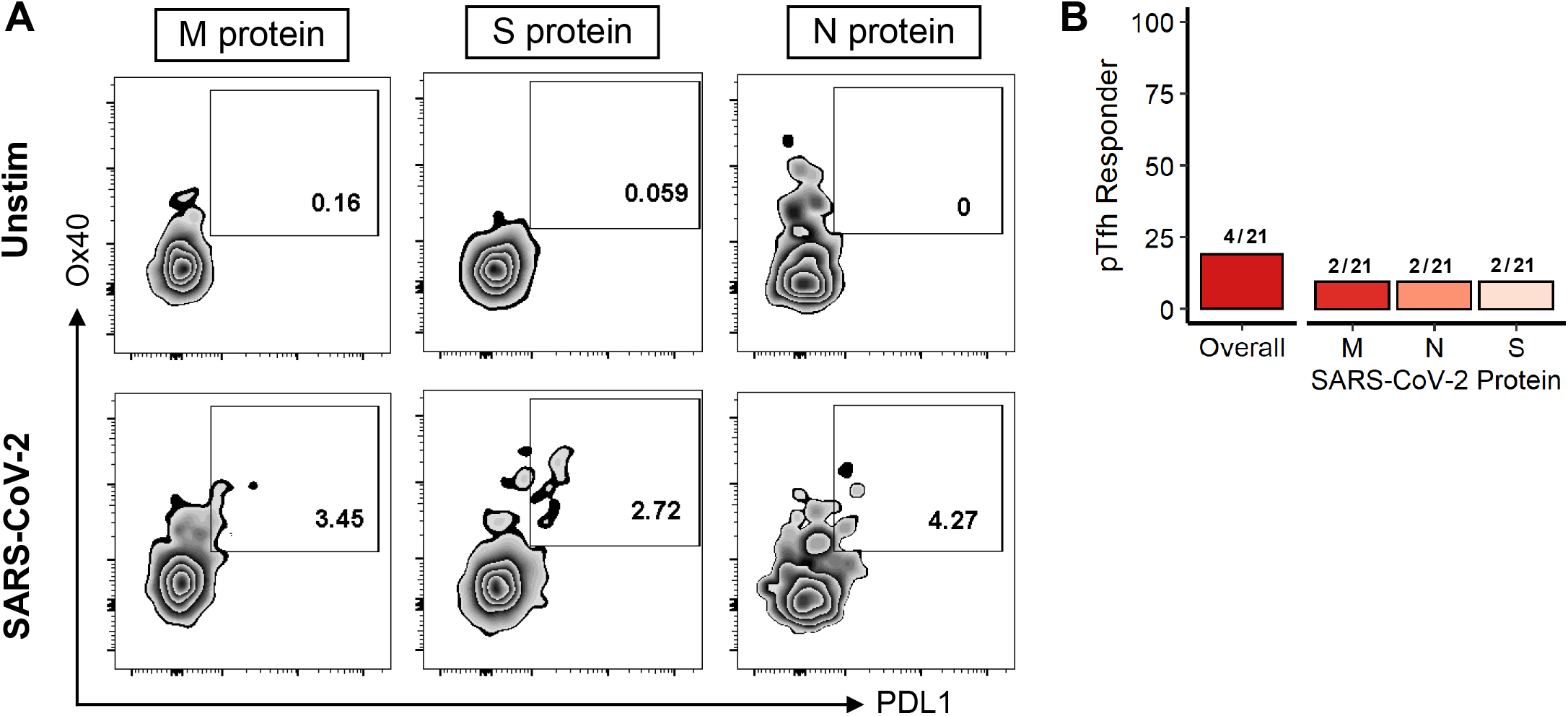
SARS-CoV-2-specific peripheral T follicular helper cells are detected at the first visit in in 4 out of 21 convalescent individuals. **(A)** Representative examples of antigen-specific pTfh (CD4^+^PD1^hi^CXCR5^+^) detected upon stimulation with SARS-CoV-2 the M, N, and S protein peptides for Visit 1 across three individuals (CR8, CR11, and CR13, respectively). Negative control of unstimulated cells shown in the top row. **(B)** Frequency of individuals mounting a positive pTfh response at their first visit to any SARS-CoV-2 protein and to the M, N, and S protein peptide pools.

Meanwhile, none of the healthy controls tested had detectable SARS-CoV-2-specific pTfh responses. This lack of pTfh responses in COVID negative individuals is not surprising, as pTfh compose a minor population of the total CD4 T cells in the blood and pTfh responses induced by other seasonal coronaviruses, if present, are likely to exist at very low, undetectable frequencies. Additionally, the fact that these responses were only detected in convalescent individuals bolsters our confidence that these pTfh responses were induced by recent SARS-CoV-2 infection and do not represent cross-reactive, memory responses.

### SARS-CoV-2-specific pTfh frequencies across the M, N, and S proteins correlate with antibody neutralization

Because pTfh are important for the development of an antibody response, we investigated whether the frequency of SARS-CoV-2-specific pTfh correlated with antibody level and neutralization at the first convalescent visit. We used two measurements of antibodies: The first was the commercially available Abbott test that detects N protein-specific IgG. The second assay measured antibody neutralization by luciferase expression and is likely a more biologically relevant metric because neutralizing antibodies have been shown to correlate with protection in preclinical studies (14, 15). For all three proteins, we see a similar level of significant correlation between the antigen-specific pTfh frequency and N protein IgG titer (**Fig 3A**). However, we find that pTfh frequencies across proteins differentially correlate with antibody neutralization (**Fig 3B)**: S protein-specific pTfh responses most strongly correlate with nAb (p < 0.0001, r = 0.75), followed by M protein-specific ones (p = 0.001, r = 0.66), and finally N protein-specific pTfh (p = 0.02, r = 0.52). To ensure these correlations were specific to SARS-CoV-2-induced responses, we quantified the frequency of total pTfh (CXCR5^+^PD1^+^). We did not see any correlation between the overall frequency of pTfh and antibody levels or neutralization (**Fig 3C**). Taken together, these data suggest that pTfh responses across SARS-CoV-2 proteins may contribute to the development of more potent nAbs.

**Figure 3:**
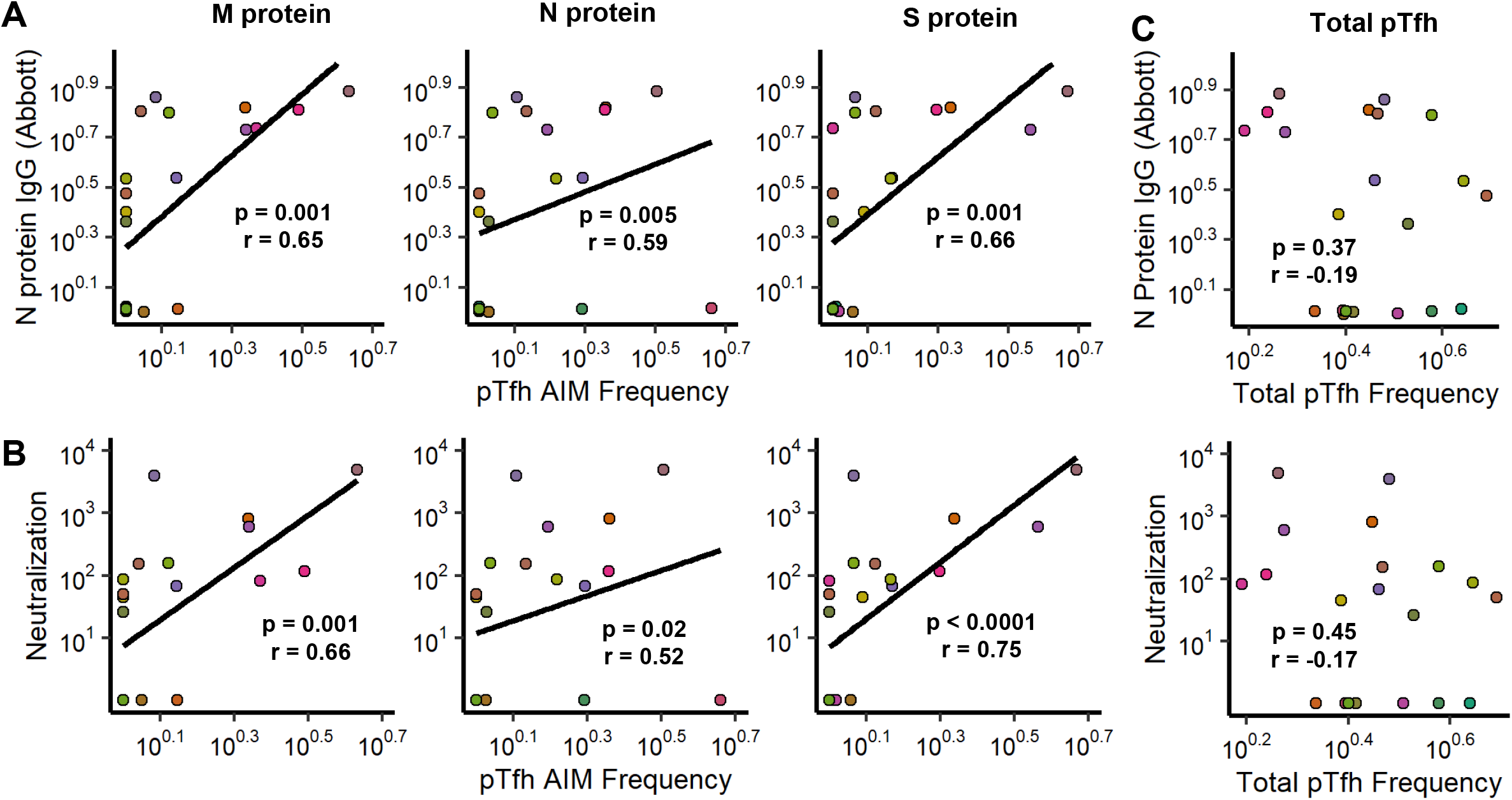
SARS-CoV-2-specific pTfh frequencies across the M, N, and S proteins correlate with antibody neutralization. **(A)** Correlations between N protein IgG titers and pTfh frequencies towards the M, N, and S proteins. **(B)** Correlations between antibody neutralization (ID50, dilution of plasma at which luminescence was reduced to 50%) and pTfh frequencies. **(C)** Correlations between the total pTfh frequency and antibody titer and neutralization. (All correlations represented by a linear regression line. Axes are transformed by log10(x+1) to allow for visualization of 0s. Statistics determined by a Spearman Correlation test. Points are colored for PTID.)

### SARS-CoV-2-specific peripheral T follicular helper responses increase over time in convalescence

To better understand the kinetics of these pTfh responses, we assessed T-cell responses in each of the convalescent individuals at a second, later visit, an average of 38 days post-symptom onset (range: 26-59 days). pTfh response frequencies detected by AIM increased from the first to second convalescent visit, where the overall pTfh responder rate went from 19% (4/21) to 43% (9/21). This increase in responses over time is most obviously observed towards the M protein where the CD4 T-cell response rate increased from 38% to 57% and the pTfh response rate increased from 10% to 33% (**Fig 4A**). Additionally, M protein-specific CD4 T-cell and pTfh response magnitudes by AIM trended up from the first to second visit (p = 0.09 and p = 0.07, respectively), while other antigen-specific subsets appeared at similar magnitudes at both timepoints (**Fig 4B**).

**Figure 4:**
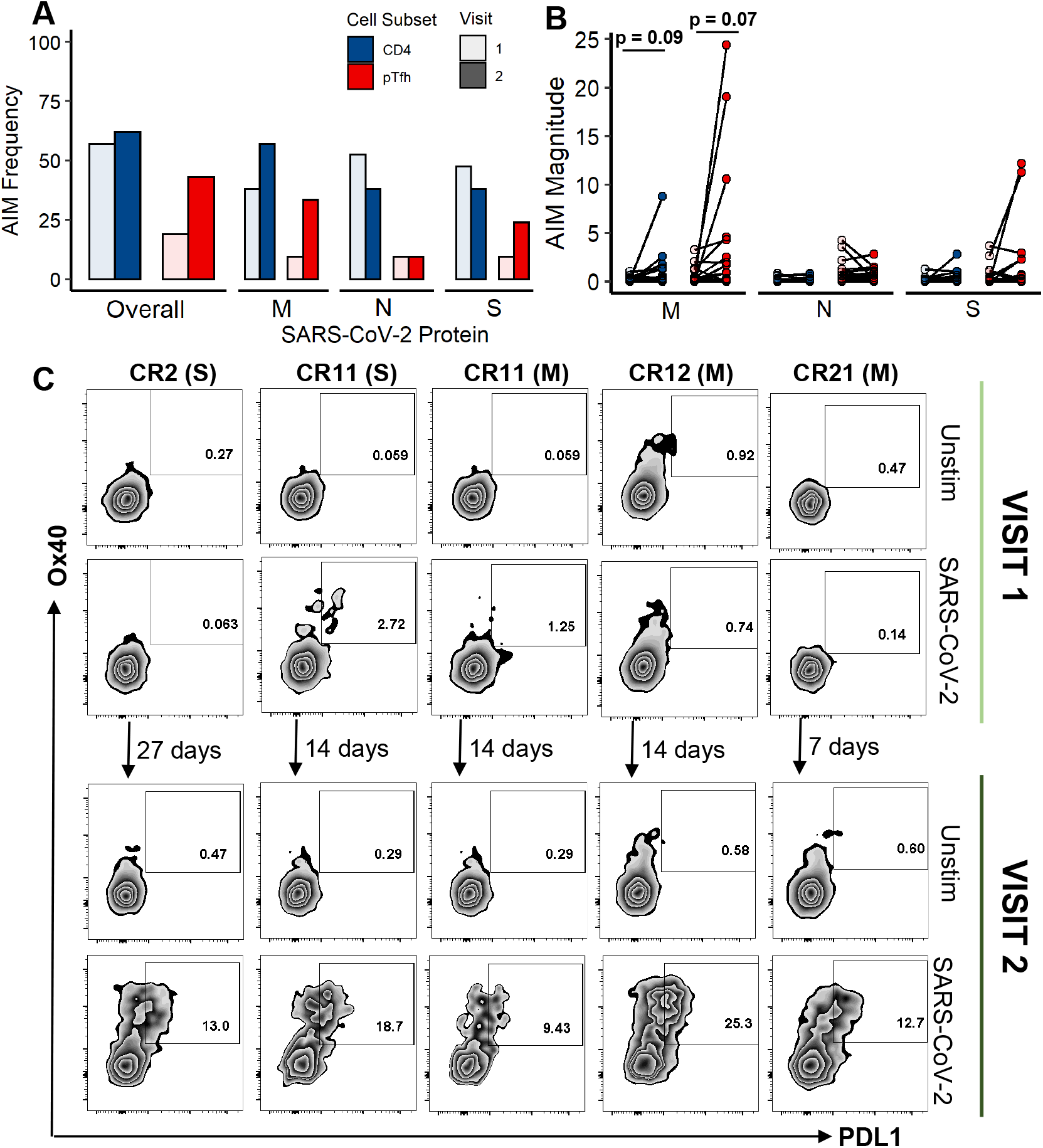
Robust SARS-COV-2-specific pTfh responses are only detected at the second convalescent visit. **(A)** Paired convalescence visit 1 and visit 2 CD4 and pTfh response magnitudes by AIM. **(B)** Paired CD4 and pTfh response magnitudes for AIM. **(C)** Flow plots for both the first (top) and second (bottom) convalescent visit of individuals where robust pTfh responses (>5%) developed. Unstimulated negative control shown for each. SARS-CoV-2 protein to which response is directed is listed next to the PTID in parentheses. (N=21, P values for magnitude comparisons determined by a paired Wilcoxon Signed-Rank Test.)

At the first visit timepoint, there were no pTfh responses with a magnitude higher than 5% frequency. Meanwhile, at the second visit, five such SARS-CoV-2-specific pTfh responses were detected in four individuals. For these four individuals, the first visit took place an average of 17 days post-symptom onset, and the second visit took place an average of 32.5 days post-symptom onset. In the case of CR21, a robust M protein-specific pTfh response arose over just seven days. These antigen stimulations are shown for both Visit 1 and Visit 2 in **Fig 4C**, and the number of days between visits is indicated between the top and bottom panels. Of these responses, only one was detected at the first visit (CR11, S protein). These responses suggest that SARS-CoV-2-specific pTfh continue to increase over time during convalescence.

## Discussion

In this study, we longitudinally examined the CD4 T-cell responses targeting the major SARS-CoV-2 structural proteins, M, N, and S, in 21 convalescent individuals by measuring the expression of activation marker and the production of effector cytokines. We found that at the first convalescent visit, antigen-specific pTfh responses could be detected against all three proteins and that the frequency of antigen-specific pTfh in these individuals correlated with nAb, albeit to varying degrees. We also found that pTfh responses increase over time in convalescence and that truly robust pTfh responses (>5% frequency) were only detected at a second, later visit.

The relative weakness of the correlation between N protein-specific pTfh frequency and antibody neutralization compared to the M and S proteins may relate back to the structure of SARS-CoV-2. Both the spike and membrane proteins have portions that are located exteriorly, while the nucleocapsid protein is found exclusively internally. Collectively, these data suggest that pTfh responses induced against different SARS-CoV-2 proteins may not be equally effective in aiding B cells and bolsters the foundation for several vaccine strategies currently in testing which only include the Spike protein. In fact, many of these vaccines have reported levels of antibodies similar to those seen in natural SARS-CoV-2 infection and mild disease, which may be a result of focusing the pTfh response on the S protein (12, 38). However, as prior studies have shown CD4 T cells across different protein specificities may contribute to nAb induction (36, 37), future studies should work to ascertain the level to which M and N protein-specific pTfh responses contribute to the formation of neutralizing antibodies. It is possible that pTfh responses across different protein specificities all play a synergistic role in the development of nAb.

Meanwhile, the observed increase in pTfh responses over time suggests that pTfh response formation may be delayed in SARS-CoV-2 infection. A study of influenza vaccination showed that pTfh responses peaked seven days after vaccine administration (25); meanwhile, a longitudinal study of pTfh in dengue virus infection found that the frequency of antigen-specific pTfh decreased from the time of acute infection (22). In comparison with these studies, it appears that pTfh response formation in SARS-Cov-2 infection continues well into convalescence as the second visit for all individuals assessed in this study occurred an average of 38 days following symptom onset. A delay in pTfh response formation could be due to the T-cell dysfunction that occurs in SARS-CoV-2 infection. Many groups have described significant T-cell dysfunction in acute SARS-CoV-2 infection (4, 5, 39), and our group has recently illustrated that this dysfunction is sustained during convalescence, even in non-hospitalized individuals (6). These high magnitude pTfh responses could also be the result of persistent antigen exposure, as several groups have reported prolonged detection of SARS-CoV-2 by PCR (40, 41). Future studies would ideally delve deeper by examining additional relevant cytokines, like IL4, IL13, and IL21, and combine activation marker and cytokine staining to allow for comprehensive functional analysis of these impressive pTfh responses arising later in convalescence.

It is also important to note that not all responses initially detected at the first visit were observed at the second visit, as illustrated by the full CD4 and pTfh response mapping by AIM and ICS (**S4A-C Fig**). When considering responses detected at either timepoint, 17/21 (81%) of individuals mounted a SARS-CoV-2-specific CD4 T-cell response by AIM (**S4D Fig**), and CD4 response were detected in 13/21 (62%) of individuals by ICS (**S4E Fig**). In fact, 43% T-cell responses detected by AIM were found at only one of the two tested timepoints. Even so, the responder frequencies detected at each visit (57% at visit 1 and 62% at visit 2, by AIM) are lower than what other recent studies have published, where SARS-CoV-2-specific T-cell responses were detected in 80-100% of individuals tested (8, 10). One reason for this is that we applied a stringent positivity criteria where responses were considered positive when three times over background and significant by fisher’s exact (p value < 0.0001), based on optimization studies conducted by the HIV Vaccine Trials Network (42). For example, for CD4 T cell responses by ICS, our responder frequency at the first visit was 48% (10/21), but if using three times the background, the CD4 responder rate is 76% (16/21). This strategy likely decreases our false positive rate but may also contribute to the discrepancy between our data and previously published studies.

These data further our understanding of CD4 T-cell responses, particularly pTfh responses, against SARS-CoV-2. Our study directly measures SARS-CoV-2-specific pTfh responses to three major structural proteins, M, N, and S. We clearly demonstrate that SARS-CoV-2-specific pTfh responses that arise early in convalescence strongly correlate with antibody neutralization and that S protein-specific responses most closely relate to antibody neutralization. But, we also show that pTfh responses against other SARS-CoV-2 proteins correlate with antibody neutralization, indicating a possible role for intrastructural help. Finally, in measuring these responses over time, we observe the emergence of several high magnitude responses more than a month following symptom onset, suggesting that pTfh response formation may be delayed in SARS-CoV-2 infection.

## Methods and Materials

### Ethics statement

All patients included in this study were adults and recruited from the University of Alabama at Birmingham (UAB) HIV care clinic, also known as the 1917 clinic, after obtaining written, informed consent and approval from the IRB-160125005 at UAB.

### Patient Samples

Cryopreserved PBMC samples for T-cell assays and plasma samples for antibody assays were acquired through the UAB COVID Enterprise Biorepository. All samples were obtained with patient consent under the appropriate IRB guidelines. Patient demographic information is shown in **Table 1**. Paired Visit 1 and Visit 2 PBMC samples from 21 individuals who had recovered from COVID-19 were assessed in this study. Clinical data from these individuals were retrieved from the Enterprise Biorepository REDCap database (43). All tested individuals were symptomatic, but none were hospitalized during the course of their illness. Symptom severity was quantified using a self-reported severity score on a scale of 1 to 3, where 1 represented no interference in daily life, 2 a moderate impact on daily life, and 3 a significant decrease in quality of life due to symptoms. A majority of individuals reported moderate severity (71%, 15/21), and a minority reported mild severity of symptoms (29%, 6/21). None reported severe symptoms. Additionally, all but two had a positive SARS-CoV-2 nasopharyngeal swab. The two individuals who did not have a PCR test completed had a known COVID contact, were symptomatic, and had detectable T-cell responses. Clinical data PBMCs from 10 healthy donors (all collected prior to the COVID-19 pandemic) were assessed for T-cell responses in parallel.

### Peptide pools

Overlapping peptides spanning the SARS-CoV-2 M, N, and S proteins (NCBI reference number MN985325.1) were designed as 20mers overlapping by 10 amino acids which has previously been shown to effectively detect CD4 T-cell responses (44, 45). Peptides were synthesized by New England Peptide in a 96-well plate format.

### Flow cytometry

For activation-induced marker staining (AIM), cells were thawed and stimulated with SARS-CoV-2 peptide pools for each of the M, N, and S proteins. An unstimulated, negative control and an SEB stimulated, positive control were included for each sample. Co-stimulatory anti-CD28 and anti-CD49d were added (BD Pharmingen). After an 18 hour incubation at 37°C, cells were washed with FACS wash (2% FBS in PBS), stained with CCR7-PercpCy5.5 at 37°C for 20 min, washed, and then stained with the following antibodies: CD4-Pe610, CD3-A780, CD8-FITC, CD14-A700, CD19-A700, Ox40-PeCy7, PDL1-PE, CXCR5-BV421, PD1-BV785, CD45RA-BV510, CD137-BV650, CD69-BUV737, and Dead cell dye-UV. Cells were then washed and fixed in 2% formaldehyde. Events were collected on a BD FACSymphony A3 within 24 hours and analyzed using FlowJo software (v10).

Intracellular staining (ICS) experiments were set up in parallel with the AIM staining experiments and performed similarly, with a few notable exceptions. CD107a-FITC was added with the co-stimulatory antibody mix; Monensin and Brefeldin A (BD Bioscience) were added after 1 hour. Cells were incubated for 12 hours in total, instead of 18. Staining was conducted in three steps: 1) Surface marker staining for 30 min at 4°C with Dead cell dye-UV, CD3-A780, CD4-BV785, CD8-V500, CD14-PercpCy5.5, and CD19-PercpCy5.5. 2) Permeabilization with CytoFix/CytoPerm solution (BD Biosciences) for 20min at 4°C. 3) Intracellular staining for 30 min at 4°C with IFNγ-A700, TNFα-PeCy7, and CD154-APC. CD154 was plotted against IFNγ. Additional details regarding the antibodies used in both the AIM and ICS assays can be found in **S1 Table**, and the gating strategies for AIM and ICS in an unstimulated, negative control are shown in **S1 Fig**.

### Antibody assays

Plasma samples from the first time point for all 21 individuals were tested for SARS-CosV-2-specific antibodies. The Abbott Architect assay was used to detect immunoglobin G (IgG) reactivity to the SARS-CoV-2 nucleocapsid protein (46). The IgG quantity is reported as a calculated index specimen/calibrator ratio, and values over 1.4 were considered positive for N protein IgG. Manufacturer-reported specificity of this assay is 99.6% (99.1%-99.9%).

Antibody neutralization assays were conducted as previously described (47). Briefly, the SARS-CoV-2 Spike (Wuhan 1, with a 19 amino acid cytoplasmic tail deletion) was pseudotyped onto an HV-1 nanoluciferase reporter backbone by co-transfection in HEC 293T cells. Pseudovirus was incubated with five-fold serial dilutions of patient plasma and then used to infect 1.5×10^4^ 293T clone 13 cells expressing ACE2. Two days post-infection, cells were washed with PBS, lysed, and nanoluciferase activity was determined according to manufacturer’s instructions (Nano-Glo® Luciferase Assay System). Luciferase activity in wells with virus and no patient plasma were set to 100%, and the dilution of plasma at which luminescence was reduced to 50% (ID50) was calculated.

### Statistical analysis

Comparisons between paired visit 1 and visit 2 magnitudes were conducted by Wilcoxon signed-rank tests. All correlations were determined by Spearman Rank tests, with the exception of Supplemental Figure 2A, where multiple measurements were plotted for each individual (across the three proteins) and therefore a generalized linear mixed effect model accounting for multiple measurements per individual was employed. In Figure 3, all axes were transformed using log10(x+1) to allow for visualization of zeros, and correlations were determined with untransformed data.

## Data Availability

Data will be made available to researcher upon request.

## Acknowledgments

We would like to acknowledge the following funding sources: internal funding from the University of Alabama at Birmingham School and Department of Medicine and F30AI140829 (SB). We would also like to thank all those who helped process samples, especially Sarah Sterrett.

## Supporting information

**Supplemental Figure 1: Flow cytometry gating strategies. (A)** Gating strategy for CD4 T cell and pTfh by activation-induced marker (AIM). **(B)** Gating strategy for CD4 T cell staining by intracellular cytokine staining.

**Supplemental Figure 2: SARS-CoV-2-reactive CD4 T cells are infrequently detected in COVID negative individuals**. Representative examples of CD4 T-cell responses detected in COVID negative individuals by upregulation of activation-induced markers **(A)** and by intracellular cytokine staining **(B)** upon stimulation by SARS-CoV-2 N protein peptide pool. Responder frequency of CD4 responses to any SARS-CoV-2 protein and to the M, N, and S proteins individually by AIM **(C)** and ICS **(D)**.

**Supplemental Figure 3: Upregulation of activation markers detected a broader range of SARS-CoV-2-specific CD4 T-cell responses. (A)** Correlation between response magnitude by AIM versus response magnitude by ICS. Statistics determined by mixed effect model accounting for multiple protein stimulations per individual. and correlation represented by linear regression line. Data transformed by log10(x+1) to allow for visualization of 0s. **(B)** Number and frequency of responses that were positive or negative by AIM and ICS. Total responses considered was 63 (3 proteins per 21 individuals).

**Supplemental Figure 4: Summary of all responses detected across two convalescent visits. (A-C)** Response summary for CD4 T cells by activation-induced marker staining, for pTfh by activation-induced marker staining, and for CD4 T cells by intracellular cytokine staining, respectively. Blue-filled cells indicate a positive response; white cells indicate a negative response. **(D)** Responder frequency by AIM across the two visits (positive at either visit) overall and to each protein. **(E)** Responder frequency by ICS across the two visits (positive at either visit).

