## supplemental table 1 for "SARS-CoV-2-specific peripheral T follicular helper cells correlate with neutralizing antibodies and increase during convalescence"

**Supplemental Table 1: Flow cytometry panels**

| AIM | | | | ICS | | | |
|---|---|---|---|---|---|---|---|
| Marker | Fluorochrome | Clone | Company | Marker | Fluorochrome | Clone | Company |
| Viability | UV | | Invitrogen | Viability | UV | | Invitrogen |
| CCR7 | PercpCy5.5 | 150503 | BD Pharminogen | CD107a | FITC | H4A3 | BD Pharminogen |
| CD4 | Pe610 | RPA-T4 | Invitrogen | CD3 | A780 | UCHT1 | Invitrogen |
| CD3 | A780 | UCHT1 | Invitrogen | CD4 | BV785 | SK3 | Biolegend |
| CD8 | FITC | SK1 | BD Biosciences | CD8 | V500 | RPA-T8 | BD Horizon |
| CD14 | A700 | M5E2 | BD Pharminogen | CD14 | PercpCy5.5 | M5E2 | BD Pharminogen |
| CD19 | A700 | HIb19 | BD Pharminogen | CD19 | PercpCy5.5 | SJ25C1 | BD Biosciences |
| Ox40 | PeCy7 | Ber-ACT35 | Biolegend | IFNγ | A700 | B27 | BD Pharminogen |
| PDL1 | PE | MIH1 | BD Pharminogen | TNFα | PeCy7 | MAb11 | BD Pharminogen |
| CXCR5 | BV421 | RF8B2 | BD Horizon | CD154 | APC | TRAP1 | BD Pharminogen |
| PD1 | BV785 | EH12.2H7 | Biolegend | | | | |
| CD45RA | BV510 | HI100 | BD Horizon | | | | |
| CD137 | BV650 | 4B4-1 | BD Horizon | | | | |
| CD69 | BUV737 | RN50 | BD Horizon | | | | |
