## supplemental figure 1 for "SARS-CoV-2-specific peripheral T follicular helper cells correlate with neutralizing antibodies and increase during convalescence"

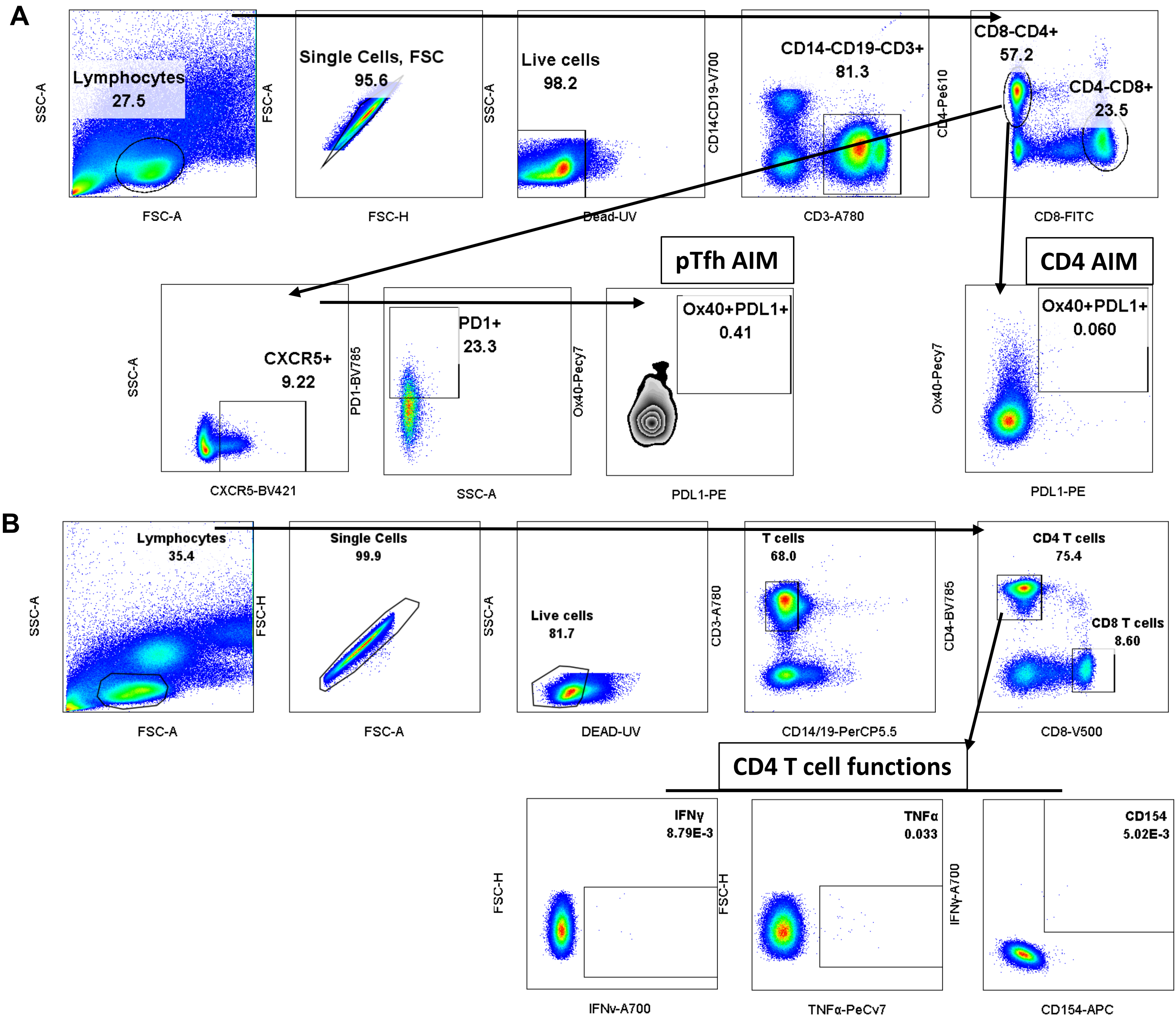


**Supplemental Figure 1: Flow cytometry gating strategies. (A)** Gating strategy for CD4 T cell and pTfh by activation-induced marker (AIM). **(B)** Gating strategy for CD4 T cell staining by intracellular cytokine staining.
