## supplemental figure 2 for "SARS-CoV-2-specific peripheral T follicular helper cells correlate with neutralizing antibodies and increase during convalescence"

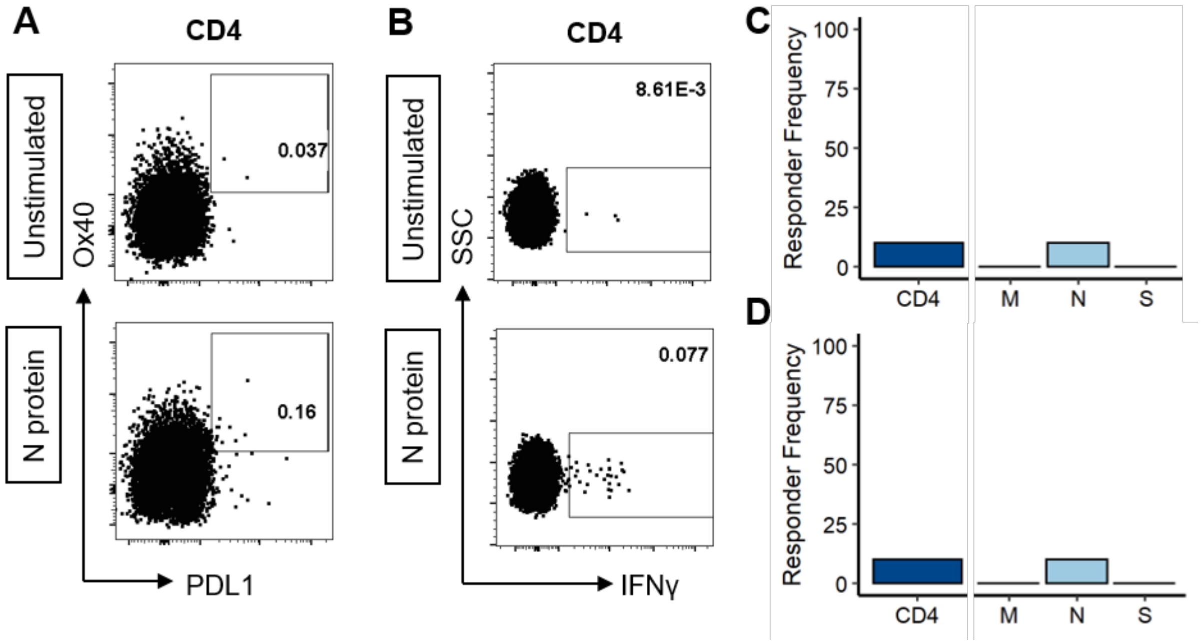


**Supplemental Figure 2: SARS-CoV-2-reactive CD4 T cells are infrequently detected in COVID negative individuals.** Representative examples of CD4 T-cell responses detected in COVID negative individuals by upregulation of activation-induced markers **(A)** and by intracellular cytokine staining **(B)** upon stimulation by SARS-CoV-2 N protein peptide pool. Responder frequency of CD4 responses to any SARS-CoV-2 protein and to the M, N, and S proteins individually by AIM **(C)** and ICS **(D)**.
