## supplemental figure 3 for "SARS-CoV-2-specific peripheral T follicular helper cells correlate with neutralizing antibodies and increase during convalescence"

**Supplemental Figure 3: Upregulation of activation markers detected a broader range of SARS-CoV-2-specific CD4 T-cell responses. (A)** Correlation between response magnitude by AIM versus response magnitude by ICS. Statistics determined by mixed effect model accounting for multiple protein stimulations per individual. and correlation represented by linear regression line. Data transformed by log10(x+1) to allow for visualization of 0s. **(B)** Number and frequency of responses that were positive or negative by AIM and ICS. Total responses considered was 63 (3 proteins per 21 individuals).

**A**

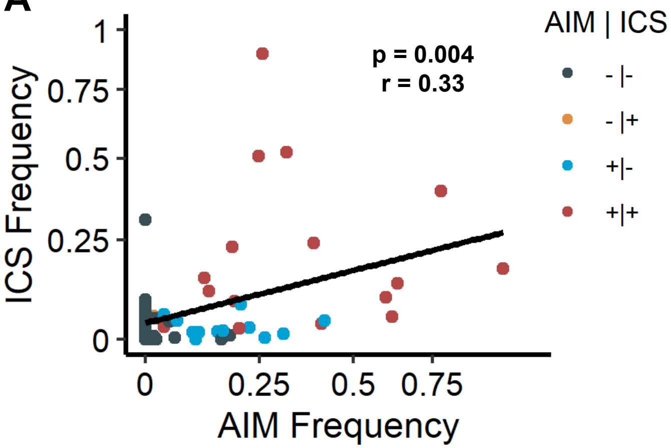


**B**

| AIM | ICS | Number | Percentage |
|---|---|---|---|
| + | + | 17 | 27% |
| + | - | 12 | 19% |
| - | + | 1 | 2% |
| - | - | 33 | 52% |
