## supplemental figure 4 for "SARS-CoV-2-specific peripheral T follicular helper cells correlate with neutralizing antibodies and increase during convalescence"

**Supplemental Figure 4: Summary of all responses detected across two convalescent visits. (A-C)** Response summary for CD4 T cells by activation-induced marker staining**,** for pTfh by activation-induced marker staining, and for CD4 T cells by intracellular cytokine staining, respectively. Blue-filled cells indicate a positive response; white cells indicate a negative response. **(D)** Responder frequency by AIM across the two visit (positive at either visit) overall and to each protein. **(E)** Responder frequency by ICS across the two visits (positive at either visit).

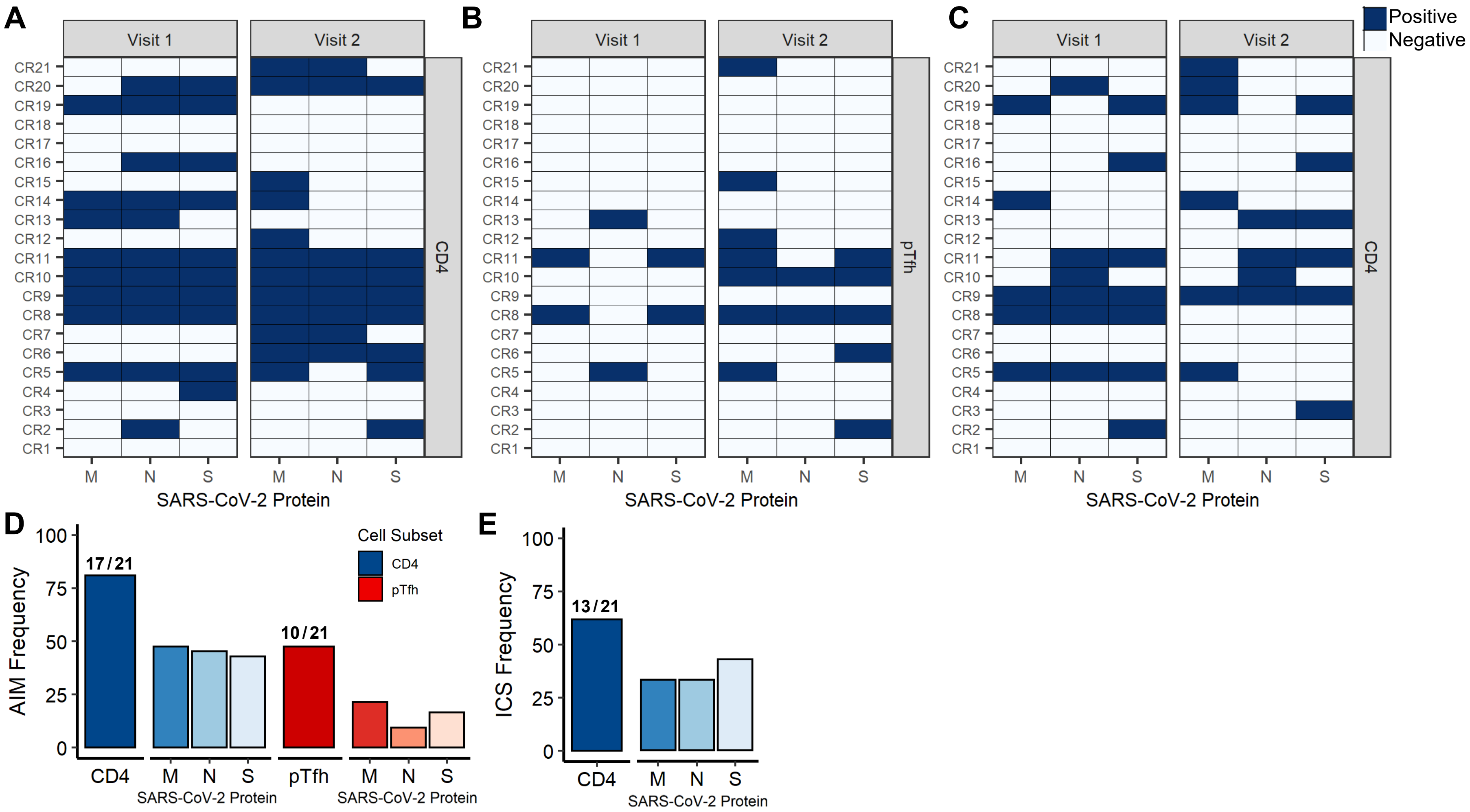
